# An approach to assess the patient benefit of demand management strategies

**DOI:** 10.1101/2023.04.16.23288617

**Authors:** Martín Yago

## Abstract

**Background:** Much of the testing performed by clinical laboratories does not translate into benefit for patients. To reduce the number of these low-value tests, laboratories use different intervention strategies, trying to adapt the analytical demand to clinical needs. The effectiveness of these interventions is usually evaluated through indicators related to the activity or cost rather than the benefit they imply for patients.

**Methods:** We have derived expressions that relate the fraction of patients tested (*R*_*t*_) and the abnormal result rate (*R*_*a*_) obtained by the requesting physician with the net benefit that the use of the test represents for both tested and untested patients.

**Results:** The behavior of physicians regarding the use of a test in each healthcare context and the effect of an intervention on this behavior can be characterized by these two parameters: *R*_*a*_ and *R*_*t*_. An increase in the value of *R*_*a*_ implies a greater net benefit for all patients attended. When the physician is selective in the use of the test, an increase in the value of *R*_*t*_ implies a greater benefit for untested patients but represents a limit to the increase in *R*_*a*_.

**Conclusions:** Interventions aimed at reducing the demand for tests should act primarily by increasing selectivity in the use of tests, increasing the benefit for the patients tested and compensating the harm that the reduction in testing entails for patients who are not.

## INTRODUCTION

Most of the critical decisions physicians make about patient care are based on information derived from laboratory tests. However, the use of tests is tremendously trivialized and terribly inefficient. Similarly to other health resources, the use of many of these tests does not translate into an obvious benefit for the patient and often represents a clear harm. The use of low-value tests produces an unnecessary consumption of resources and puts the patient at risk due to over-investigation, overdiagnosis, and overtreatment (1). For this reason, laboratories have used different intervention strategies to try to adapt the demand for testing to clinical needs, which has been called demand management (2).

Although low value testing should be viewed primarily as a risk to the patient rather than a cost issue, the success of these interventions is typically purely evaluated on laboratory indicators (e.g. percentage reduction in tests performed or the money saved by the laboratory) rather than the proportion of tests used inappropriately or the benefit that the intervention represents for patients.

Many of the published studies (3) on the use of laboratory tests establish the inappropriate use of a test by comparing it to what it is recommended by clinical guidelines through a clinical audit procedure. Although this approach is aligned with the normative theory of rationality postulated by evidence-based medicine, it is costly and has several limitations. On the one hand, there is often a shortfall of appropriate recommendations for a given testing indication and there are many recommendations with little evidence or simply inconsistent with each other (4). On the other hand, given the diversity of healthcare settings and clinical presentations, it is sometimes a challenge in practice to objectively establish whether a test is appropriate or not. Moreover, the extrapolation of the recommendations of the clinical guidelines to the particularities of each patient is precisely a cause of the uncertainty that drives the variability in clinical practice (5). The inappropriate use of a test is essentially characterized by suboptimal patient selection by the requesting physician. According to Bayes’ theorem, a low selectivity will result in a low pretest probability and consequently a low diagnostic value of the test (6). Since physicians who are more selective in the use of tests are expected to obtain a higher proportion of abnormal results, the abnormal result rate has been used as a parameter to directly assess the relative selectivity of laboratory testing without the need to resort to chart audits in order to determine the pretest probability of a positive result (7,8).

The objective of this work has been to develop a new, more practical, objective and patient-centered approach to assess the effects of interventions carried out for demand management. Using the concept of expected utility and assuming that the inappropriate use of a test is basically due to the lack of selectivity of the requesting physician, we have derived expressions that relate the testing behavior of the physician (indexed by the rate of abnormal results obtained) to the benefit that testing represents both for the tested patients and for those who are not. Based on these relationships and using a simple example, a procedure is described to determine how a demand management intervention translates to the use of tests by the requesting physicians and to what extent it benefits the patients.

## METHODS

### Model description

The results of this study have been obtained using a model based on the concept of expected utility, which is frequently used in normative theories for rational decision making. It is assumed that the use of a test is beneficial for a patient if it allows an accurate diagnosis and is followed by the appropriate treatment. Conversely, the use of the test will be detrimental to the patient when misdiagnosed and mistreated due to the diagnostic limitations of the test.

The expected utility of using the test is the weighted average of the utilities that each of all possible outcomes (true positives and negatives, and false positives and negatives) have for the patient, where each utility is a measure of the degree to which that result is preferred by the patient with respect to the rest. The utility of each outcome is weighted according to the probability that using the test will lead to that outcome. A more detailed description of the model is given in the Appendix in the Supplemental Material that accompanies the online version of this article.

Similarly, the expected utility for a patient who has not been tested will be the weighted average of the utilities of a true negative (the patient does not have the disease), and a false negative (the patient actually does).

The net benefit for the tested patients (*NB*_*T*_) is the difference between the expected utility of receiving the test and that of not having received it. *NB*_*T*_ is linearly related to the prevalence of the investigated disease or clinical condition among the tested patients (*P*_*T*_) according to the following equation:

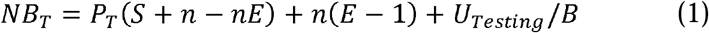

where *S* and *E* are the clinical sensitivity and specificity of the test respectively, and *n* is the ratio between the cost of a false positive result and the benefit of a true positive result (*B*) valued by the patient according to their preferences. *U*_*Testing*_ represents the utility of carrying out the test (economic cost or other type of inconvenience).

Similarly, the net benefit for the patient who has not been tested *NB*_*U*_ will be the difference between the utility of not receiving the test and the utility of having received it. *NB*_*U*_ is linearly related to the prevalence of the investigated disease among the untested patients, *P*_*U*_:

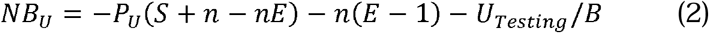

A net benefit with a positive value indicates a benefit to the patient and a negative value indicates a cost or harm. More detail regarding the derivations of the above equations is given in the Appendix.

### Selectivity using the test

The *P*_*T*_ and *P*_*U*_ values are determined by the physician’s selectivity when making the request. A physician is selective using a test when the pre-test probability for the patients for whom the test is requested is greater than the one of the total number of patients being attended. The more selective the physician, the higher the *P*_*T*_ and the lower the *P*_*U*_ and therefore a greater benefit for both the tested and untested patients. The most selective physicians are also characterized by obtaining a higher abnormal results rate, *R*_*a*_, according to the well-known Rogan and Gladen formula (9):

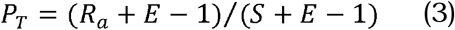

Substituting Eq. 3 into 1, and rearranging, it can be verified that there is a linear relationship between *R*_*a*_ and *NB*_*T*_ :

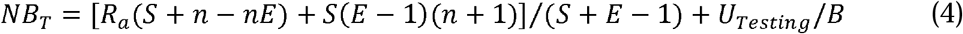

This means that the professional who uses a test more selectively will obtain a higher rate of positive results and a greater benefit for the patients tested.

However, *P*_*U*_ does not depend only on the selectivity of the requesting physician, but also on the fraction of patients attended that are tested, *R*_*t*_. The relationship between *P*_*U*_ and *R*_*t*_ is given by the following equation:

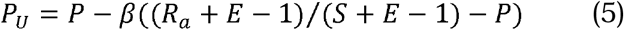

where *P* is the prevalence of the disease in the population of patients attended and *β = R*_*t*_*/(1 - R*_*t*_), that is, the ratio between the fraction of tested and untested patients (see Appendix for derivation).

According to this equation, if a physician is selective using the test, *P*_*U*_ decreases as *β* increases, because the more patients are selectively tested, the lower the prevalence of disease among untested patients and thus the higher *NB*_*U*_. The variation of *P*_*U*_ as a function of *β* will be more pronounced the higher the value of *R*_*a*_. On the contrary, if the physician preferentially requests the test for patients who do not have the disease, the second term of the equation will be negative and *P*_*U*_ will increase when β does, to the detriment of the untested patients. If the physician is non-selective *P*_*T*_ *= P*, and *P*_*U*_ will be independent of *R*_*t*_.

### Effect of an intervention on the net benefit to patients

If for some reason, such as a demand management intervention, a change occurs that may affect the selectivity of clinicians in using a test or the fraction of patients under their care who receive the test, the variation in the net benefit that this change represents for tested patients (Δ*NB*_*T*_) and non-tested (Δ*NB*_*U*_) will be given by the following formulas:

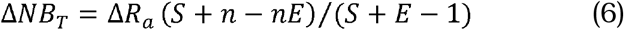

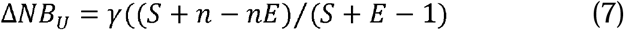

where Δ*R*_*a*_ *= R*_*a1*_ *- R*_*a0*_, and *R*_*a0*_ and *R*_*a1*_ are respectively the *R*_*a*_ values before and after the intervention. *β*_*0*_ and *β*_*1*_ are also the *β* values before and after the intervention; Δ*β = β*_*1*_ *- β*_*0*_ ; *γ = R*_*a1*_ *β*_*1*_ *- R*_*a0*_*β*_*0*_ *- m*Δ*β* and *m = P*(*S+ E - 1*) - (*E- 1*).

It can be shown (see Eq. 16 in the Appendix) that *m* is the expected value for *R*_*a*_ in the absence of selectivity (i.e. when the test is used randomly without considering the patient’s pretest probability or when all patients seen by the physician are tested). The model assumes that neither the prevalence of the disease studied in the population of patients attended nor the diagnostic properties of the test change because of the intervention.

Given that Δ*NB*_*T*_ is directly proportional to Δ*R*_*a*_ and Δ*NB*_*U*_ is directly proportional to *γ*, these two parameters allow to determine whether the effect of a demand management intervention for a test has or has not been beneficial for patients, both for those who undergo the test and for those who do not. These parameters also indicate the magnitude of the effect of the intervention, both globally and at the level of the individual requesting physicians, and allow an analysis of the behavior for each of them considering their selectivity and the rate of use of the test in the patients they care for.

The estimation of Δ*R*_*a*_ and *γ* for each physician requires knowing the number of tests requested, the proportion of abnormal results obtained, and the number of patients seen during a given period, data that is normally accessible to laboratories. The estimation of *γ* also requires knowing the value of *m*, a more difficult parameter to obtain. However, given that *m* is the value that *R*_*a*_ takes when *R*_*t*_ *=1* and that there is a linear dependence between *R*_*a*_ and *1/R*_*t*_, it is possible to estimate *m* using a linear regression.

### Application example

The application of these concepts to assess the effect of a simple intervention to improve the use of serum γ-glutamyltransferase (GGT) measurement by primary care physicians in our health department is illustrated below. The intervention basically consisted of removing this test from a liver profile containing ALT, total bilirubin, ALP, and GGT; and performing GGT only when ALP results were above reference values. The use of the test by each physician during the year prior to the intervention was compared with that of the following year. Of a total of 83 physicians in the department, 17 were excluded from the comparison because they did not show significant healthcare activity (less than 500 consultations/year) during either of these two periods. The estimation of *m* was performed by Kendall–Theil Sen Siegel nonparametric linear regression using the “mblm” package in R (10). A file containing the raw data and the calculations performed is included as Supplemental Material.

## RESULTS

The equations obtained from the model used in this study indicate that there is a linear relationship between the net benefit that the use of a test entails for the patients tested (*NB*_*T*_) and the abnormal results rate obtained by the requesting physician (*R*_*a*_), a measure of the physician’s selectivity, i.e. their ability to distinguish the patient who may benefit from the test from the one who will not. Interestingly, this relationship is independent of the prevalence of the disease investigated among the patients seen by each physician, so that two physicians with the same *R*_*a*_ value provide the same benefit to the patients they test, even though they serve populations with different prevalence. In this case, however, the physician with a lower population prevalence will have to be more selective to achieve equal benefit.

Patients who are attended by the physician but not tested also benefit when the selectivity of the requesting physician is high, although in this case the net benefit of the intervention for these patients (*NB*_*U*_) also depends on the fraction of patients on whom the physician uses the test, *R*_*t*_. For a given level of selectivity (i.e., for a constant *R*_*a*_) the net benefit for the untested patient is greater the higher the value of *R*_*t*_, provided that the test is used with some selectivity and not in a completely random manner.

Therefore, the behavior of the physicians w.r.t. the use of a diagnostic test, from the point of view of the benefit that this represents for the patients they attend and in a certain healthcare context, can be characterized by the corresponding values of *R*_*a*_ and *R*_*t*_ for each and every physician.

Fig. 1 shows the values of these two parameters for the use of the GGT by 66 primary care physicians before and after the intervention carried out to improve the use of this test described above. It is observed that there is an inverse relationship between the two parameters. As *R*_*t*_ exceeds the prevalence of disease among the patients seen, inevitably a larger number of patients who do not have the disease will be tested, limiting the maximum selectivity that can be achieved. In this sense, *R*_*t*_ represents an upper bound for the value of *R*_*a*_. The extreme case would occur when *R*_*t*_ *=1* (all patients seen by the physician are tested); in which the selective use of the test is obviously impossible. The value of *R*_*a*_ that would be obtained if the request for the test was performed randomly (in a non-selective way) *m*, has been estimated from these data, by linear regression, as 0.120 (95% CI [0.114, 0.132]).

**Fig. 1.**
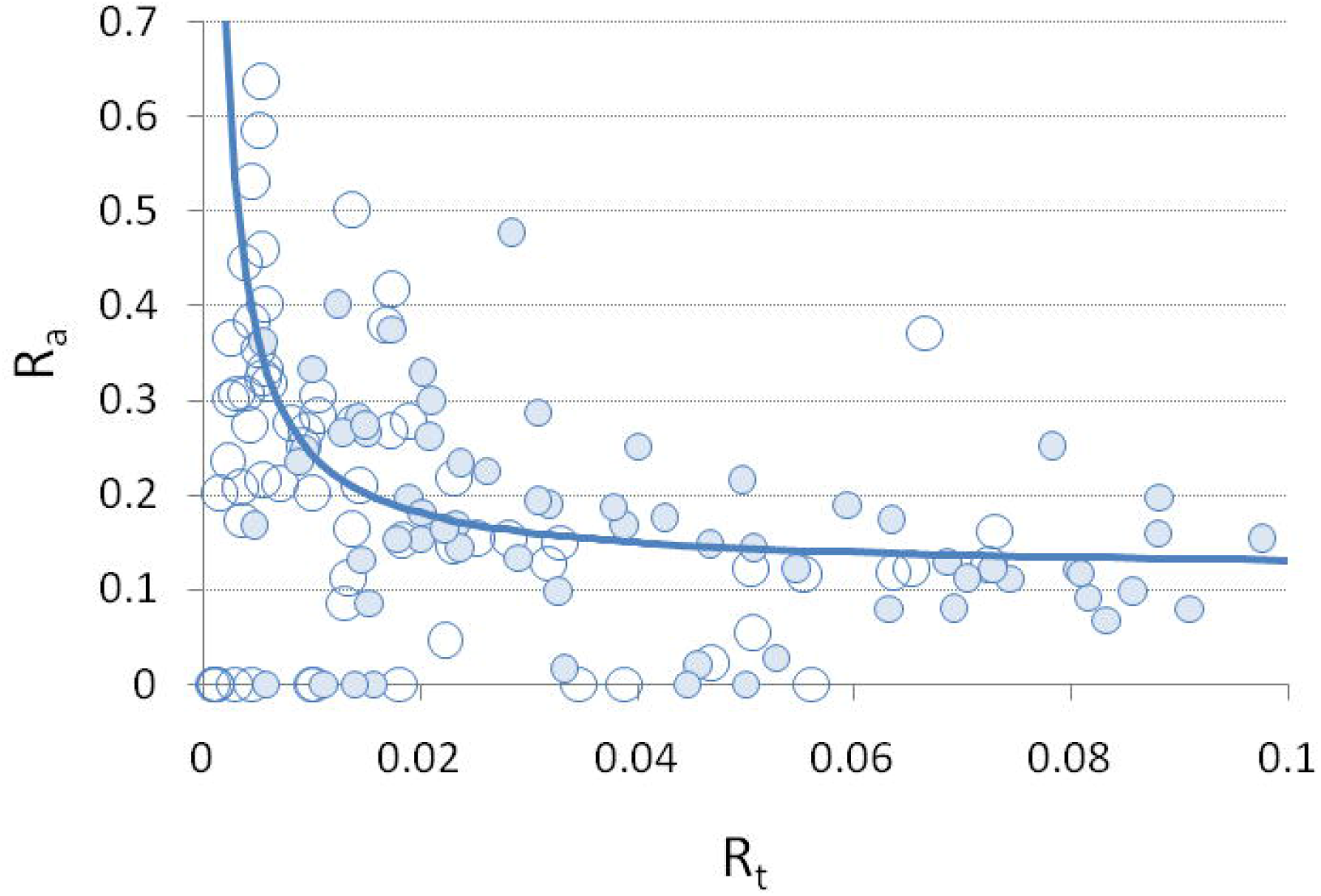
The fraction of patients tested (*R*_*t*_) versus the abnormal result rate (*R*_*a*_) for 66 physicians before (open circles) and after (dark circles) the intervention to improve GGT utilization. The regression line is shown.

Overall, a 47% decrease (from 8,923 to 4,677 tests/year) was observed in the total number of GGT tests requested by the physicians studied during the year following the intervention compared to the previous year. Although the intervention could be considered a success from the point of view of reducing demand, one should ask whether it was really beneficial for the patients attended by these physicians.

Fig. 2 represents the observed change in behavior in the use of the test by each physician coinciding with the intervention. The change in *R*_*a*_, Δ*R*_*a*_, a parameter that is directly proportional to the change in net benefit for tested patients, Δ*NB*_*T*_, is plotted against *γ* a parameter that is directly proportional to the change in net benefit for untested patients, Δ*NB*_*U*_ (see equations 6 and 7). A positive value of these parameters indicates a greater net benefit for patients after the intervention.

**Fig. 2.**
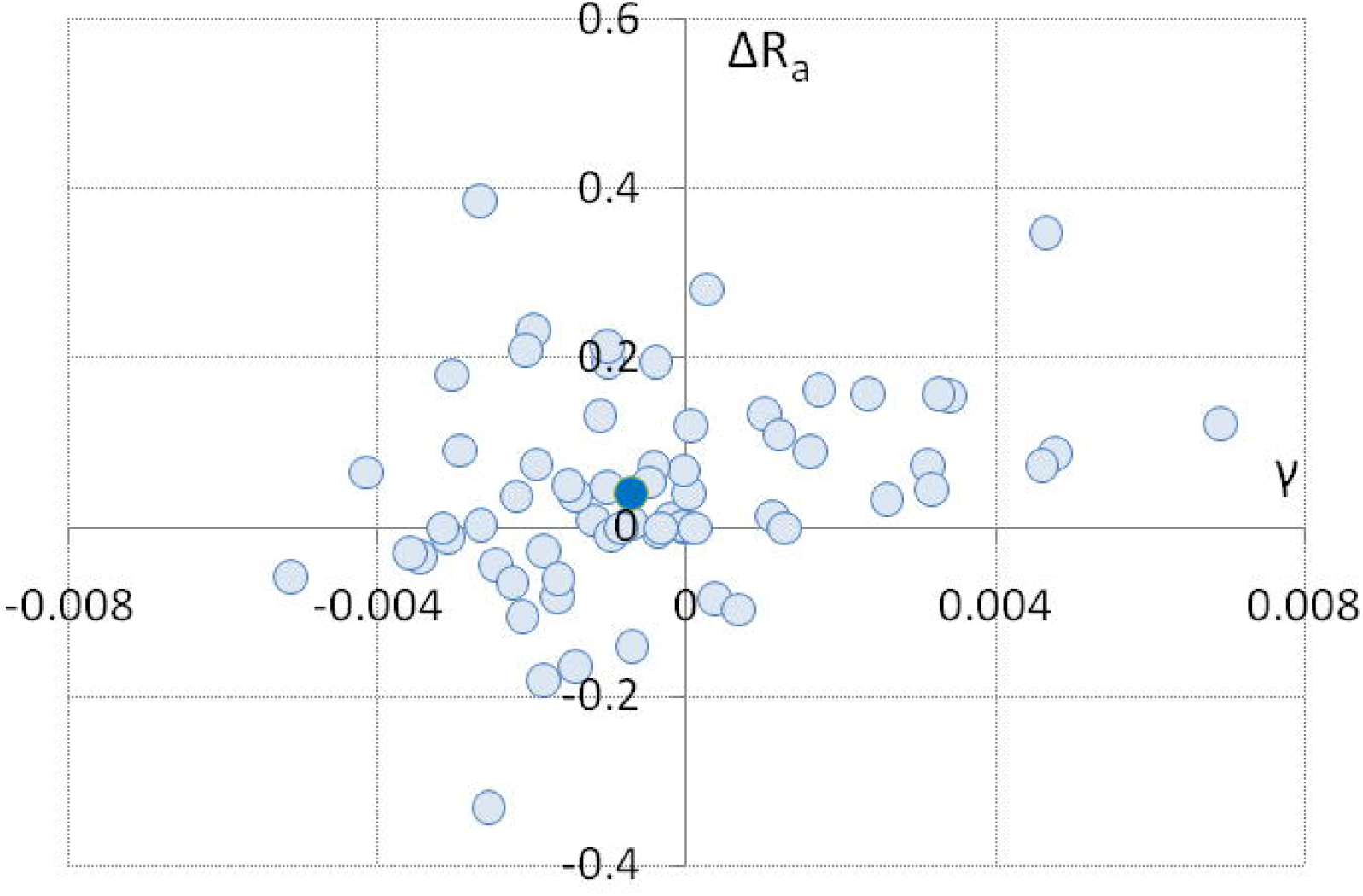
The change in the abnormal result rate against the gamma parameter for 66 physicians undergoing the intervention to improve GGT utilization. The dark dot indicates the median for the two parameters.

The median of the Δ*R*_*a*_ values was 0.0377 (95% CI [0.0197, 0.0795]), a statistically significant increase (Wilcoxon signed rank test, P=0.002). This means that physicians tend to use the test more selectively after the intervention, leading to an increase in the net benefit for the patients tested.

The median of the *γ* values was -0.00071 (95% CI [-0.00107, 0.00011]), although the decrease is not significant (Wilcoxon signed rank test, P=0.1). This means that the decrease in demand for the test observed after the intervention is excessive to achieve a beneficial effect for untested patients. Although an increase in selectivity in the use of the test also increases the net benefit for untested patients, a decrease in the proportion of patients who receive the test has the opposite effect and can nullify it, as in this case.

On the other hand, the physicians who have most improved the use of the test after the intervention are those who use it most intensively. The physicians who after the intervention have increased the net benefit for all their patients, tested and untested, are the 21 that appear in the upper right quadrant of Fig. 2. The median *R*_*t*_ for this group of physicians before the intervention was almost three times that of their peers (0.069 vs. 0.024; Wilcoxon rank sum test, P<0.001). This effect occurs because the increase in net benefit for untested patients when selectivity increases is proportionally greater the greater the use of the test (see discussion of Eq. 5 in the Materials and Methods section).

## DISCUSSION

The application of strategies to adapt analytical demand to clinical needs, usually called demand management, has become widespread in clinical laboratories in recent decades, which have considered it part of their professional responsibility and integrated it as part of their culture. Although these strategies should be aimed at improving clinical outcomes, indicators used to assess the progress and success of this type of interventions usually have little relation to the harms and benefits they have for the patients. In addition, they may have objectivity problems or be inconvenient for routine use and they are usually global measures that do not take into account the inter-individual variability characteristic of the requesting physicians for the analysis and interpretation of the results of the intervention.

The procedure proposed in this paper is based on two indicators that are usually available in healthcare activity records: the abnormal result rate and the proportion of patients seen by each physician who are tested. An increase in the value of the first indicator for the physician who uses the test supposes an increase in the net benefit both for the patient who is tested and for the one who is not. Increases in the second indicator may represent a net benefit for untested patients, but only when the use of the test is selective, and acts as a limiting factor on the magnitude of possible increases in selectivity. For these reasons, interventions aimed at reducing the demand for tests should act primarily by increasing selectivity in the use of tests (and not reducing demand as usual), thus increasing the benefit for the patients tested and compensating the harm that the reduction in testing entails for patients who are not. It should be noted, however, that the reduction in the demand for low-value tests that allows an increase in selectivity also indirectly represents a benefit for all patients by improving equity in access to health resources, reducing the environmental impact of health care activity and allowing the sustainability of health systems based on social solidarity.

According to this approach, a higher abnormal result rate always indicates a better use of the test, although this does not mean that the value that a negative result can have in excluding a diagnosis or clinical condition is ignored. Even when a test is used for this purpose, there must always be a reason to suspect the clinical condition to be excluded, so a physician who appropriately orders the test will tend to obtain a higher proportion of abnormal results than one who uses it randomly or less selectively. This is the case, for example, of the systematic verification of the homeostatic stability of inpatients, probably the main cause of overuse of laboratory tests in the hospital setting.

The proposed method improves the objectivity of the evaluation of the effects of interventions for demand management. For this purpose, objective indicators of activity are used instead of indicators based on the proportion of tests used inappropriately established by the opinion of experts taking the clinical guidelines as a reference. The need to establish absolutely whether a test use is appropriate in each individual patient is avoided by using relative measures of change in test use rates and abnormal result rates obtained by each individual clinician.

However, a drawback of this approach is the need to assume that the preferences regarding the use of the test and the prevalence of the investigated clinical condition among patients seen by different physicians are the same. Although the latter is not necessary to establish the benefit of the intervention for the tested patients, it is necessary to establish that of the non-tested ones.

On the other hand, this procedure characterizes the behavior of each requesting physician and allows to know which aspects of the use of the test (selectivity or request rate) can be improved in each one of them. This information can be helpful in deciding what type of strategy or combination of strategies to use. For example, the use of reflex algorithms is appropriate to increase selectivity, the design of request forms may be appropriate to modify the request rate, and the use of order alerts may influence both aspects. However, the interventional strategy that is effective for one physician may not be for another, and its widespread use may even be counterproductive. For example, in the design of interventions aimed at reducing demand based on feedback strategies, it is necessary to decide whether to tackle the approach of the most overusing physician or rather focus on the information that is presented and on how it is presented. Detailed data on the behavior of each requesting physicians can help make these types of decisions and, more importantly, can help determine which of them may be the most effective. This opens the door to performing more individualized interventions, aimed exclusively at improving the desired aspects and restricted to physicians who really need it.

## Supporting information

raw data

## Data Availability

All data produced in the present work are contained in the manuscript

## Nonstandard abbreviations

*R*_*a*_: abnormal result rate
*R*_*t*_: fraction of tested patients
*NB*_*T*_: net benefit for the tested patient
*NB*_*U*_: net benefit for the untested patient
*P*_*T*_: prevalence of the investigated disease among the tested patients
*P*_*U*_: prevalence of the investigated disease among the untested patients

## Notes

### Competing Interest Statement

The authors have declared no competing interest.

### Funding Statement

This study did not receive any funding

