## Appendix for "An approach to assess the patient benefit of demand management strategies"

**Model Description**

Let us suppose that the following situation arises: a physician taking care of a patient considers carrying out a certain laboratory test, thinking that this may be useful to investigate and establish a treatment for a suspected disease, syndrome, or clinical condition. We will also assume that the test result is interpreted dichotomously (normal or abnormal) and that an abnormal result (positive) translates into treatment initiation. The treatment can have negative consequences for the patient, but if the diagnosis is correct, the patient can benefit from it.

Here the term "treatment" should be understood in its broadest sense: it may consist of starting a pharmacological treatment, modifying an existing one, establishing a waiting period or simply subjecting the patient to more diagnostic tests.

With all, the test is not perfect and can produce false-positive and false-negative results that put the patient at risk of receiving inappropriate treatment or not receiving it when it is really needed.

Figure S1 portrays a model with all the possible outcomes for the described situation. Each of the possible outcomes is coupled with a probability of occurrence and a utility. A utility is a numerical value for a health benefit, harm, or monetary cost, measured on a common scale. A positive value for utility indicates a benefit and a negative value indicates a cost or harm.

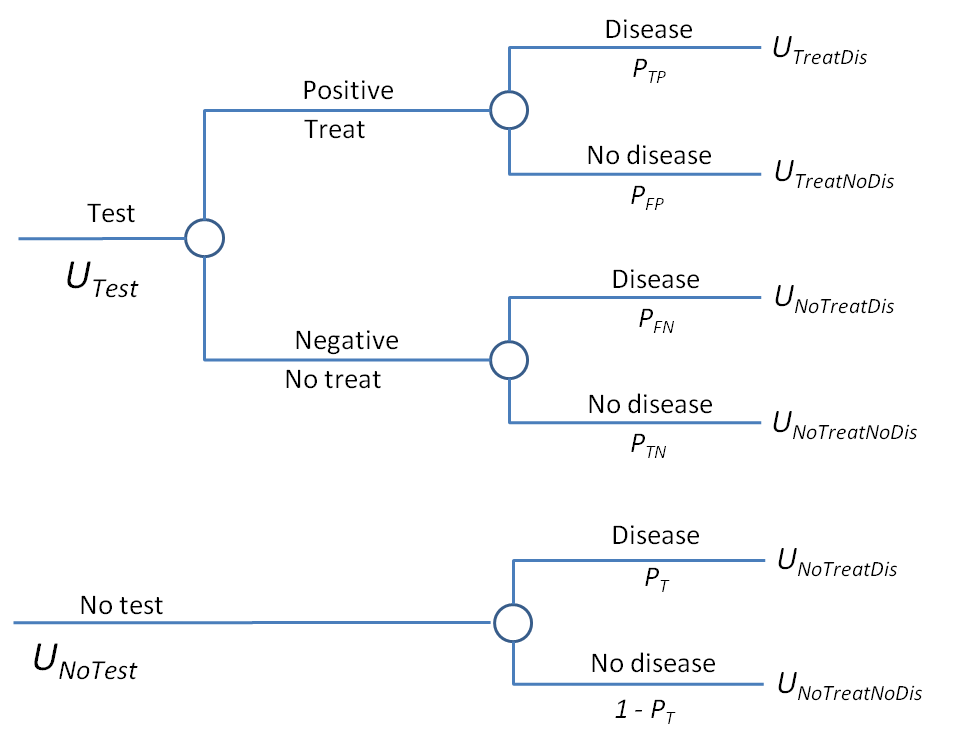

Figure S1. Utilities and probabilities of all possible outcomes associated with the use of a test in a population with a prevalence of disease $P_{T}$.

Five utilities are associated with carrying out the test:

- $U_{TreatDis}$ - the utility of treating a person who has the disease
- $U_{NoTreatDis}$ - the utility of not treating a person who has the disease,
- $U_{TreatNoDis}$ - the utility of treating a person who does not have the disease
- $U_{NoTreatNoDis}$ - the utility of not treating a person who does not have the disease and
- $U_{Testing}$ - the utility (harm or cost) associated with testing.

The expected utility of performing the test, $U_{Test}$ , is the average of the utilities weighted by their probabilities of occurrence:

$U_{Test}= P_{TP}U_{TreatDis}+P_{FP}U_{TreatNoDis}+P_{FN}U_{NoTreatDis}+P_{TN}U_{NoTreatNoDis}+U_{Testing}$ (1)

where $P_{TP}$ is the probability that a result is a true positive, $P_{FP}$ is the probability of a false positive, $P_{FN}$ is the probability of a false negative and $P_{TN}$ the probability of a true negative. All these probabilities add up to one.

Similarly, the expected utility of not performing the test (and thus not treating the patient) is:

$U_{NoTest}= P_{T}U_{NoTreatDis}+(1-P_{T})U_{NoTreatNoDis}$	(2)

where $P_{T}$ is the prevalence of the investigated disease or clinical condition among the tested patients.

**Net benefit for the patients tested**

The benefit that performing the test represents for the patients is the difference between the expected utility of performing the test and the counterfactual, that is, the expected utility of not testing:

$U_{Test}-U_{NoTest}=P_{TP}U_{TreatDis}+P_{FP}U_{TreatNoDis}+{P_{FN}U}_{NoTreatDis}+P_{TN}U_{NoTreatNoDis}-P_{T}U_{NoTreatDis}-(1-P_{T})U_{NoTreatNoDis}+U_{Testing}$	(3)

Substituting	$P_{FN}=P_{T}-P_{TP}$ and $P_{TN}=1-P_{T}-P_{FP}$ in Eq. 3 and rearranging terms:

$U_{Test}-U_{NoTest}=P_{TP}\left( U_{TreatDis}-U_{NoTreatDis} \right)-P_{FP}(U_{NoTreatNoDis}{-U}_{TreatNoDis})+U_{Testing}$	(4)

where the difference $B=U_{TreatDis}-U_{NoTreatDis}$ represents the benefit of treatment for a person who has the disease and $C=U_{NoTreatNoDis}-U_{TreatNoDis}$ represents the cost of treating a person when it is not necessary. Dividing Eq. 4 by $B$ to express the benefit of performing the test relative to the benefit of treatment for a person with the disease, we obtain the "net benefit" for the person tested, ${NB}_{T}$, a concept widely used for evaluating the clinical usefulness of diagnostic tests and prediction models (1,2):

${NB}_{T}=P_{TP}-P_{FP}(C/B) +U_{Testing}/B$	(5)

And finally, substituting $P_{TP}=P_{T}S$ and $P_{FP}=\left( 1-P_{T} \right)(1-E)$ in Eq. 5, we obtain:

${NB}_{T}=P_{T}\left( S+n-nE \right)-n{\left( 1-E \right)+U}_{Testing}/B$	(6)

where$S$ is the clinical sensitivity of the test, $E$ is the clinical specificity and $n=C/B$. In patient-centered care, $n$ represents the patient's preferences. The more the patient values the benefit of being treated appropriately ($B$) in relation to the harm of being treated unduly ($C$), the lower the value of $n$ and the greater the net benefit of performing the test.

**Net benefit for the untested patients**

Suppose now that the physician treating the patient has not deemed it appropriate to perform the test. In this case, the benefit to the patient of not performing the test can be calculated as the difference between the expected utility of not performing the test and the expected utility of performing it.

${{U_{NoTest}-U}_{Test}=P}_{U}U_{NoTreatDis}+\left( 1-P_{U} \right)U_{NoTreatNoDis}-(P_{TP}U_{TreatDis}+P_{FP}U_{TreatNoDis}+{P_{FN}U}_{NoTreatDis}+P_{TN}U_{NoTreatNoDis})-U_{Testing}/B$	(7)

where $P_{U}$ is the prevalence of the investigated disease or clinical condition among the untested patients. And therefore, the net benefit for the untested patient can be calculated as:

${NB}_{U}={-P}_{U}\left( S+n-nE \right)+n{\left( 1-E \right)-U}_{Testing}/B$	(8)

The higher the value of $n$, the greater the benefit for the patient who is not tested.

**Selectivity using the test**

In general, not all patients have the same probability of receiving a certain test. By virtue of their selectivity in the use of the test, health professionals can classify their patients into two groups: one of them with a greater probability of having the condition under investigation who will be tested, and another group less likely to have the condition, where the use of the test seems unnecessary and therefore will not be tested.

This selectivity, $\alpha$, can be expressed as the ratio between the probability of performing the test on a person who really has the condition, $P_{Dis}$, and the probability of performing it on someone who really does not have it,$P_{NoDis}$:

$\alpha={P_{Dis}}/{P_{NoDis}}$	(9)

Normally, only a fraction $R_{t}$ of the patients seen by a professional are tested, which can be calculated as the fraction of tested patients who have the disease plus the fraction of tested patients who do not have it:

$R_{t}=PP_{Dis}+(1-P)P_{NoDis}$	(10)

where $P$ is the prevalence of the disease among the patients attended by the professional and $R_{t}$is the ratio between the number of people who receive the test and the total number of people attended.

The prevalence of the disease among the patients tested, $P_{T}$, will be equal to the ratio between the fraction of tested patients who have the disease and the fraction of patients tested:

$P_{T}={{PP_{Dis}}/{R_{t}}=\alpha P}/{(\alpha P-P+1)}$	(11)

$P_{T}$ will be equal to the prevalence of the condition in the patients, $P$, when the professional requesting the test does not show selectivity and uses the test purely at random ($\alpha=1$). Note that the relationship between $P_{T}$ and $\alpha$ is hyperbolic: the higher $P_{T}$ is, the greater the increase in $\alpha$ must be in order to achieve the same increase in $P_{T}$.

In case the professional was able to identify and perform the test on all patients with the disease, $P_{Dis}=1$ would hold and according to Eq. 11,$P/R_{t}$ would be the maximum value that $P_{T}$ can take for a given value of $R_{t}$. Therefore, the value of $R_{t}$ sets an upper bound for $P_{T}$ that cannot be exceeded.

For example, if the prevalence of the disease in the population of patients treated is 0.1 and the physician requests the test to a 20% of all the patients being treated, the pre-test probability in the patients tested can never exceed 0.5 (0.1/0.2=0.5).

The most selective professionals are those who use the test with a higher pre-test probability, $P_{T}$, and therefore obtain a higher rate of abnormal results, $R_{a}$. The relationship between these two parameters is given by the well-known Rogan and Gladen equation (3):

$P_{T}={{(R}_{a}+E-1)}/{(S+E-1)}$	(12)

Similarly, the prevalence of disease among untested patients, $P_{U}$, will be equal to the ratio between the fraction of untested patients who have the disease and the fraction of untested patients:

$P_{U}={P(1-P_{Dis})}/{(1-R_{t})}$	(13)

Substituting Eq. 11 into 13 we get:

$P_{U}=P-\beta(P_{T}-P)$	(14)

where $\beta={R_{t}}/{(1-R_{t})}$, that is, the ratio between the fractions of tested and untested patients; and substituting Eq. 12 in Eq. 14 we obtain:

$P_{U}=P-\beta({{(R}_{a}+E-1)}/{(S+E-1)}-P)$	(15)

which indicates that $P_{U}$ is lower the greater $R_{a}$ and the greater $\beta$, although the latter is only true when the professional shows some selectivity (i.e. when $P_{T}>P$) and the second term of the equation15 is positive.

Finally, combining equations 12 and 14 it is shown that there is an inverse linear relationship between $R_{a}$ and $\beta$:

$R_{a}=(1/\beta)\left( S+E-1 \right)\left( P-P_{U} \right)+m$	(16)

where $m=P(S+E-1)-\left( E-1 \right)$, is the value of $R_{a}$ in the absence of selectivity (i.e. when $P_{U}=P$).

**Effect of an intervention on the net benefit for patients**

Suppose now that there is a change of some kind in the care setting or a demand management intervention is performed that changes the way clinicians use the same test but without changing the cost or harm associated with performing the test or patient preferences. The intervention, however, may change the selectivity with which professionals using the test or the frequency with which they use it.

The change in net benefit as a consequence of the intervention for the patients who are tested will be the difference between the net benefit after, ${NB}_{T1}$, and before, ${NB}_{T0}$, the intervention. From Eq. 6:

${\Delta NB}_{T}={{NB}_{T1}-{NB}_{T0}=\Delta P}_{T}\left( S+n-nE \right)$	(17)

where ${\Delta P}_{T}=P_{T1}-P_{T0}$ and $P_{T1}$ and $P_{T0}$ are the prevalence of the disease in tested patients after and before the intervention, respectively.

Similarly, the change in net benefit as a consequence of the intervention for patients who are not tested will be the difference between the net benefit after, ${NB}_{U1}$, and before, ${NB}_{U0}$, of the intervention. From Eq. 8:

${\Delta NB}_{U}={{NB}_{U1}-{NB}_{U0}=-\Delta P}_{U}\left( S+n-nE \right)$	(18)

where ${\Delta P}_{U}=P_{U1}-P_{U0}$ , and $P_{U1}$ and $P_{U0}$ are the prevalence of the disease in untested patients after and before the intervention, respectively.

Any change that occurs in *α* because of a demand management intervention or other circumstance will produce a change in $R_{a}$ and consequently a proportional change in ${\Delta NB}_{T}$. Substituting Eq. 12 into 17:

${\Delta NB}_{T}={\Delta R}_{a}\left( S+n-nE \right)/{(S+E-1)}$	(19)

where ${\Delta R}_{a}=R_{a1}-R_{a0}$, and $R_{a0}$ and $R_{a1}$ are the $R_{a}$ values before and after the intervention, respectively.

Similarly, any change in $\alpha$ or $R_{t}$ will cause a change in ${NB}_{U}$. Substituting Eq. 14 into 18:

${\Delta NB}_{U}=\left[ \left( P_{T1}-P \right)\beta_{1}-\left( P_{T0}-P \right)\beta_{0} \right](S+n-nE)$	(20)

and substituting Eq. 12 into 20:

${\Delta NB}_{U}=\gamma({S+n-nE)}/{(S+E-1})$	(21)

where $\gamma=R_{a1}\beta_{1}-R_{a0}\beta_{0}-m\Delta\beta$ and $m=P(S+E-1)-\left( E-1 \right)$.

According to this equation, a decrease in the use of the test does not necessarily lead to a decrease in ${NB}_{U}$ because it can be compensated by an increase in selectivity. In order to keep ${NB}_{U}$ invariant by setting $\gamma=0$, the following equation establishes the change that must occur in $R_{a}$to compensate for a change in $\beta$:

$\beta_{1}={\beta_{0}(m-R_{a0})}/{(m-R_{a1})}$	(22)
